# Brain tumor MRI classification and identification using an image classification model via Convolutional Neural Networks

**DOI:** 10.1101/2024.09.13.23299832

**Authors:** Naitik Mohanty, Morteza Sarmadi

## Abstract

Malignant brain tumors are generally classified to be extremely aggressive and often can be fatal when not met with immediate action. Glioblastoma Multiforme is the most common type of malignant tumor found in the brain and is extremely aggressive. For this reason, advanced detection of malignant brain tumors is necessary for optimal mitigation. Conversely, the classification of tumors during Medical Resonance Imaging can be difficult due to bodily movements resulting in the movement of the tumor. The movement of the tumor can disrupt targeted radiotherapy and can also, at times, result in treatments about radiotherapy damaging healthy areas of the brain rather than areas of the tumor. This study proposes a novel deep learning system that can identify tumors from MRI images; which can be helpful for the case of early detection, as well as being able to track tumors during active imaging; resulting in higher efficiency with targeted radiotherapy. This is done utilizing Convolutional Neural Networks (CNNs) created via deep learning frameworks. With the image identification of tumors; 97% accuracy was achieved with optimization. The tumor-classification deep learning system achieved an accuracy of 98%. Further testing is required for optimization; with this optimization, higher accuracy can be reached.

## 1 INTRODUCTION

Malignant brain tumors are characterized by their aggressive nature and the imminent threat they pose to human life. Among these formidable foes, Glioblastoma Multiforme (GBM) stands as the most common type of malignant brain tumor, notorious for its ruthless progression and relentless assault on the brain. A timely and accurate response to the presence of such tumors is of paramount importance (Pereira et.al). The delicate nature of the human brain, coupled with the dynamic challenges it presents, has long prompted the pursuit of innovative solutions in the field of medical imaging and diagnosis. In this pursuit, one pioneering solution has emerged – a novel deep learning system that leverages Convolutional Neural Networks (CNNs) to identify and track brain tumors from MRI images, offering ease of early detection, more efficient treatment, and enhanced precision in targeted radiotherapy.

Glioblastoma Multiforme, often referred to simply as GBM, poses a formidable challenge in the medical world. It is aggressive, relentless, and commonly lethal when not addressed promptly and effectively. GBM is characterized by its rapid growth and infiltrative nature, making it notoriously difficult to treat. To combat this, a multifaceted approach is required, involving early detection, precise diagnosis, and effective treatment strategies (Zreik et.al). Medical Resonance Imaging (MRI) has proven to be a vital tool in the diagnosis and monitoring of brain tumors. However, it is not without its challenges. The brain’s constant movement within the skull, influenced by factors such as respiration and heartbeat, can result in the displacement of tumors during imaging, posing a significant obstacle to accurate diagnosis and classification. This issue is not only detrimental to the patient but also hampers the efficacy of targeted radiotherapy, a critical treatment modality for brain tumors. In some unfortunate instances, the misalignment caused by tumor movement during treatment may lead to healthy brain tissue being inadvertently damaged, instead of the tumor itself.

Recognizing the urgent need to address these challenges and improve the accuracy and efficiency of brain tumor diagnosis and classification, this study introduces an innovative deep-learning system. This system harnesses Convolutional Neural Networks, which have been tested in image analysis tasks, and integrates them into medical imaging (Pereira et.al). The primary objective of this study is twofold. First, to enhance the early detection of malignant brain tumors by identifying them from MRI images with a high degree of accuracy. Second, to develop a tumor classification system that can adapt to the dynamic nature of the brain, ensuring precise monitoring during active imaging and treatment. Early detection can significantly improve patient outcomes by allowing for prompt intervention, while accurate classification ensures that the tumor’s location remains known throughout treatment, maximizing the precision of targeted radiotherapy and minimizing collateral damage to healthy brain tissue. The research team achieved an accuracy rate of 97% with careful optimization, demonstrating the system’s potential to reliably identify brain tumors from MRI images. This level of accuracy is a significant step forward in the quest for early diagnosis, and it offers hope to patients and medical practitioners alike.

The tumor classification system, while still undergoing optimization, has achieved a commendable accuracy rate of 98%. This represents a substantial advancement in the field of real-time tumor monitoring during medical imaging. Although further testing and optimization are required, the potential benefits are abundantly clear. Enhanced classification will enable healthcare professionals to adapt treatment strategies in real time, ensuring that the tumor remains precisely within the crosshairs of targeted radiotherapy.

## 2 METHODS

### 2.1 Construction of Tumor Identification Module

#### 2.1.1 Data Curation and Cleaning forTumor Identification Module

For the tumor identification module, we first used a dataset released by Columbia University with 253 MRI scans. The set contains 137 scans with no tumors and 116 scans with tumors. The dataset used provides T1-weighted imaging which helps identify tumor cell clustering. Figure 1 depicts the imaging modality gained from the dataset.

**Figure 1:**
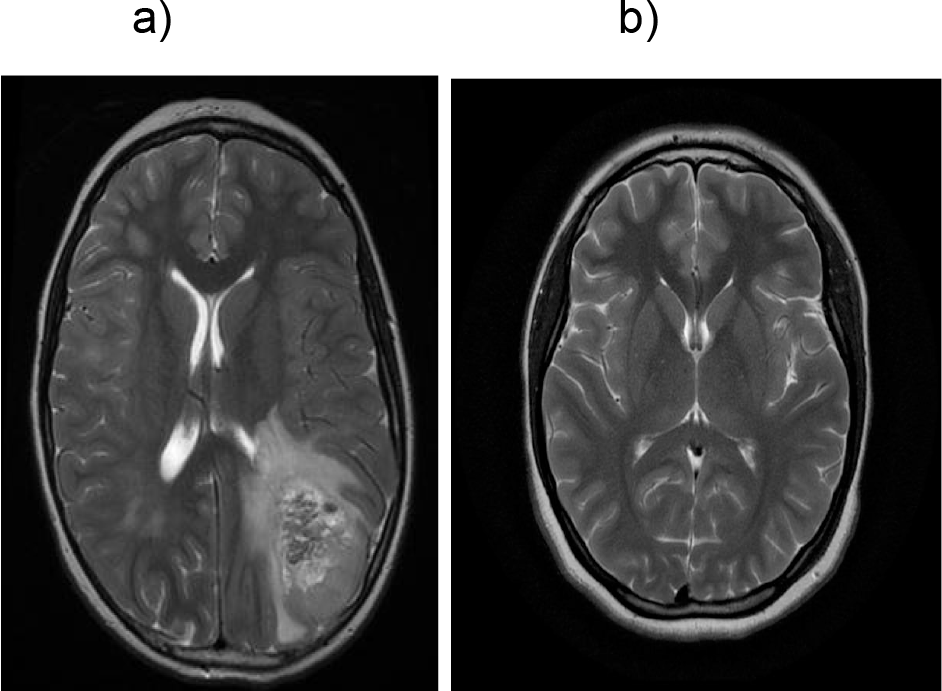
T1Gd imaging gained from 2 patients. One with imaging indicating a tumor and one without an imaging indication of a tumor. a)T1GD with tumor; b)T1Gd without tumor

For the data curation; a variety of methods were used. First, the image was cropped only to include parts of the brain; excluding blank spaces for a more accurate model with a shorter run-time. This was done by cropping the scan’s extreme top, left, bottom, and right portions. To better understand the cropping process; figure 2 is shown to depict a cropped image compared to the original image.

**Figure 2:**
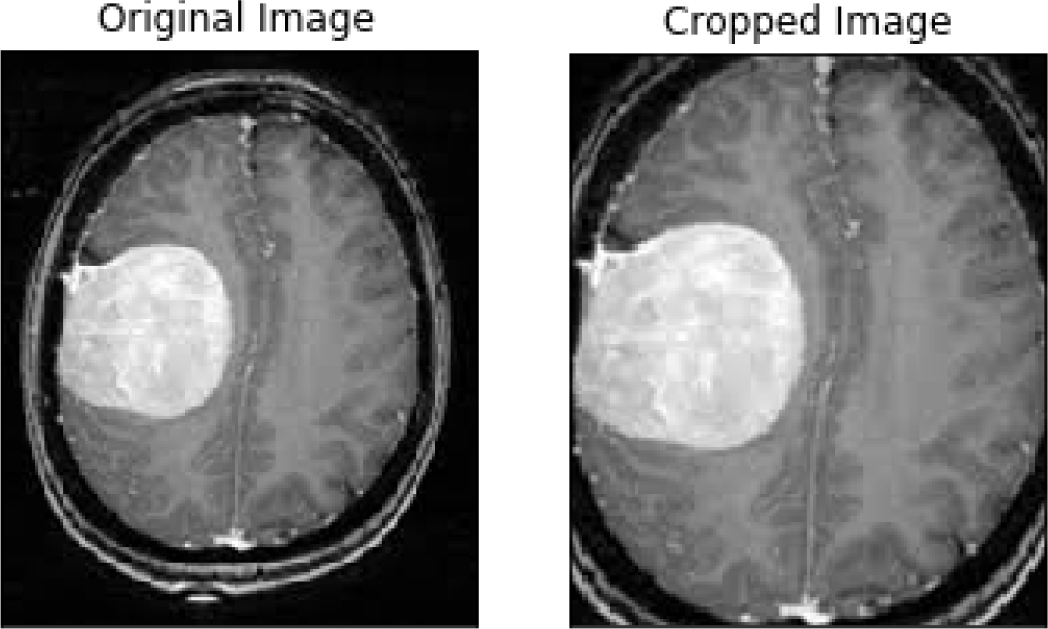
Cropped V.S. Original Image. Original Image, side by side to image cropped to exclude the extreme top, left, bottom, and right portions.

The training dataset was then loaded. Which would include 50 MRI imaging scans without a tumor and 50 MRI scans with a tumor. We decided to split the dataset into 3 portions; approximately consisting of these 3 categories: 70% for network training, 15% for validation, and 15% for testing. This would help ensure adequate accuracy and set appropriate boundaries for dataset utilization.

#### 2.1.2 CNN Model Creation

We used a convolutional neural network to create the image identification section. Specifically, we used a sequential model; where each layer only connects to the directly previous and directly following layers. The model begins with an input layer specifying an input shape of (224, 224, 3), which is common for color images (224×224 pixels with 3 color channels). The model then adds two convolutional layers, each followed by a max-pooling layer. The first convolutional layer has 80 filters with a kernel size of (3,3), a ReLU activation function, and “valid” padding. The max-pooling layer reduces the spatial dimensions by a factor of 2. The second convolutional layer consists of 64 filters with similar settings. These layers are designed to extract features from the input image data. The flattened layer converts the 2D feature maps into a 1D vector, passing through a series of densely connected layers. The model includes two hidden layers with 500 units and ReLU activation, followed by dropout layers with a 30% dropout rate to prevent overfitting. Finally, an output layer with a single unit and a sigmoid activation function indicates a binary classification task. The model is compiled using the Adam optimizer and binary cross-entropy loss, and accuracy is used as a metric for training and evaluation.

This sequential model is designed for image classification tasks and utilizes convolutional layers to learn hierarchical features from the input images. The max-pooling layers reduce the spatial dimensions, making the model computationally efficient while retaining important information. The fully connected layers at the end of the network provide a means for higher-level feature combinations and decision-making. Dropout layers are introduced to reduce the risk of overfitting by randomly setting a fraction of input units to zero during training. The output layer with a sigmoid activation function suggests that this model is meant for binary classification problems, where it aims to output a probability of the input belonging to a particular class. Overall, this architecture can be used for tasks like image classification, where the goal is to classify input images into one of two categories.

#### 2.1.3 Model Training

The training phase of the deep learning model uses the Keras library. It is training the previously defined sequential model on a dataset. The ‘model. fit’ method is used to train the model. It takes several arguments, including the training data (’x_train_scaled’ and ‘y_train’) and the number of training epochs, which is set to 10 in this case. During each epoch, the model will iteratively update its internal parameters to improve its performance on the training data. The ‘validation_data’ argument is set to a tuple containing the validation data (’x_test_scaled’ and ‘y_test’), allowing the model’s performance to be evaluated on a separate dataset during training to monitor its generalization ability.

The training process involves iteratively forwarding the training data through the model, calculating the predicted outputs, computing the loss between the predicted outputs and the true labels, and then using this loss to update the model’s internal parameters (weights and biases) via backpropagation and the specified optimization algorithm (Adam in this case). The training process is repeated for the specified number of epochs, to minimize the binary cross-entropy loss as specified in the model’s compilation. By the end of the training, the model’s performance on both the training data and validation data will be stored in the ‘history’ variable, which can be used to analyze the training progress, check for overfitting, and make decisions about model improvements or early stopping based on metrics such as accuracy and loss. This code represents a fundamental step in the development of machine learning models, where the model learns to make predictions based on the provided data and labels.

#### 2.1.4 Model Testing and Results

Following the training of the network, the model is asked to print out the predictions for what images it believes to be “Tumor” or “No Tumor”. As seen below in figure 3.

**Figure 3:**
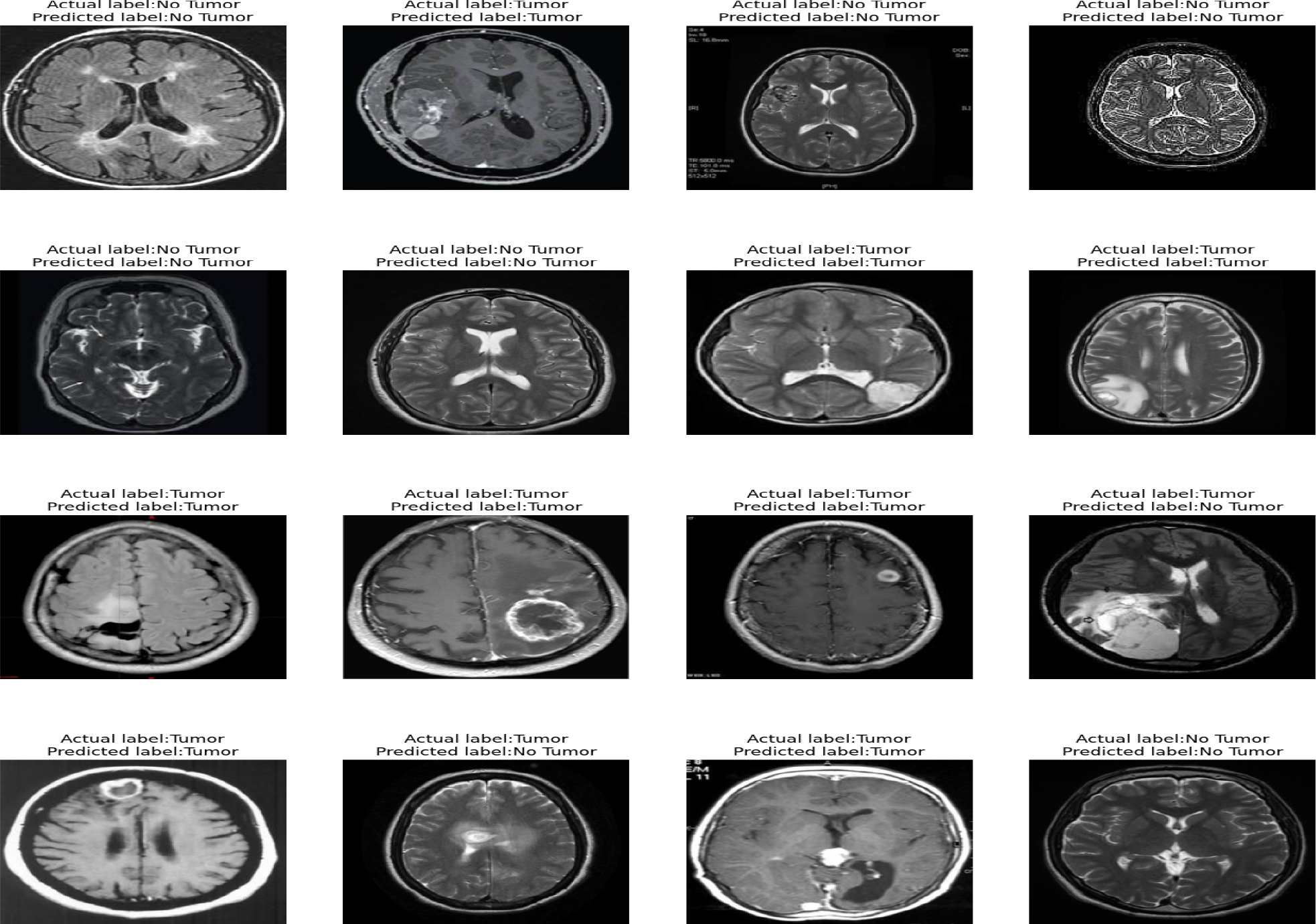
CNN model prediction. First printout of predictions from CNN model classifying images with “Tumor” or “No tumor”.

In the first testing, the model achieved an accuracy of 87% and slightly leaned toward correctly classifying images as “No Tumor” rather than “Tumor”, showing 88% accuracy for images with no tumor and 86% for images with tumors; taking 120 seconds and 931 ms. This can be remedied by changing the learning rate of the CNN model. The learning rate was changed from 10^−1^ to 10^−3^. This would greatly enhance the accuracy to 97% with an 11% increase from the original testing. The model is again asked to print out the predictions of what images it believes are “Tumor” or “No Tumor”. As seen in Figure 4.

**Figure 4:**
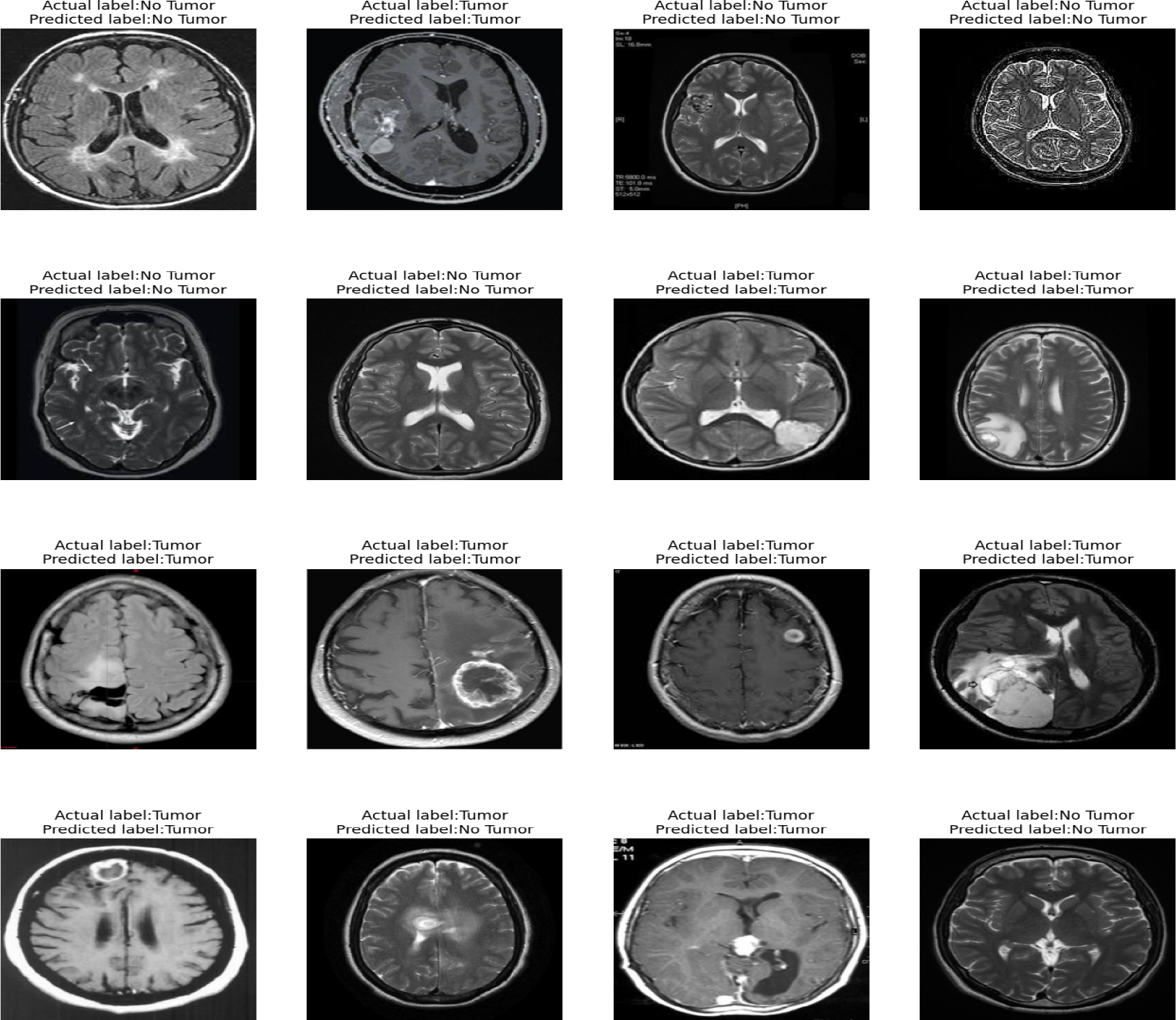
CNN model prediction. Updated printout of predictions from CNN model classifying images with “Tumor” or “No tumor with custom learning rate

The sensitivity was found to be 96.4, the precision was found to be 89, and the specificity was found to be 94.5. Graph showing accuracy with epoch number is shown in *Figure 5*

**Figure 5:**
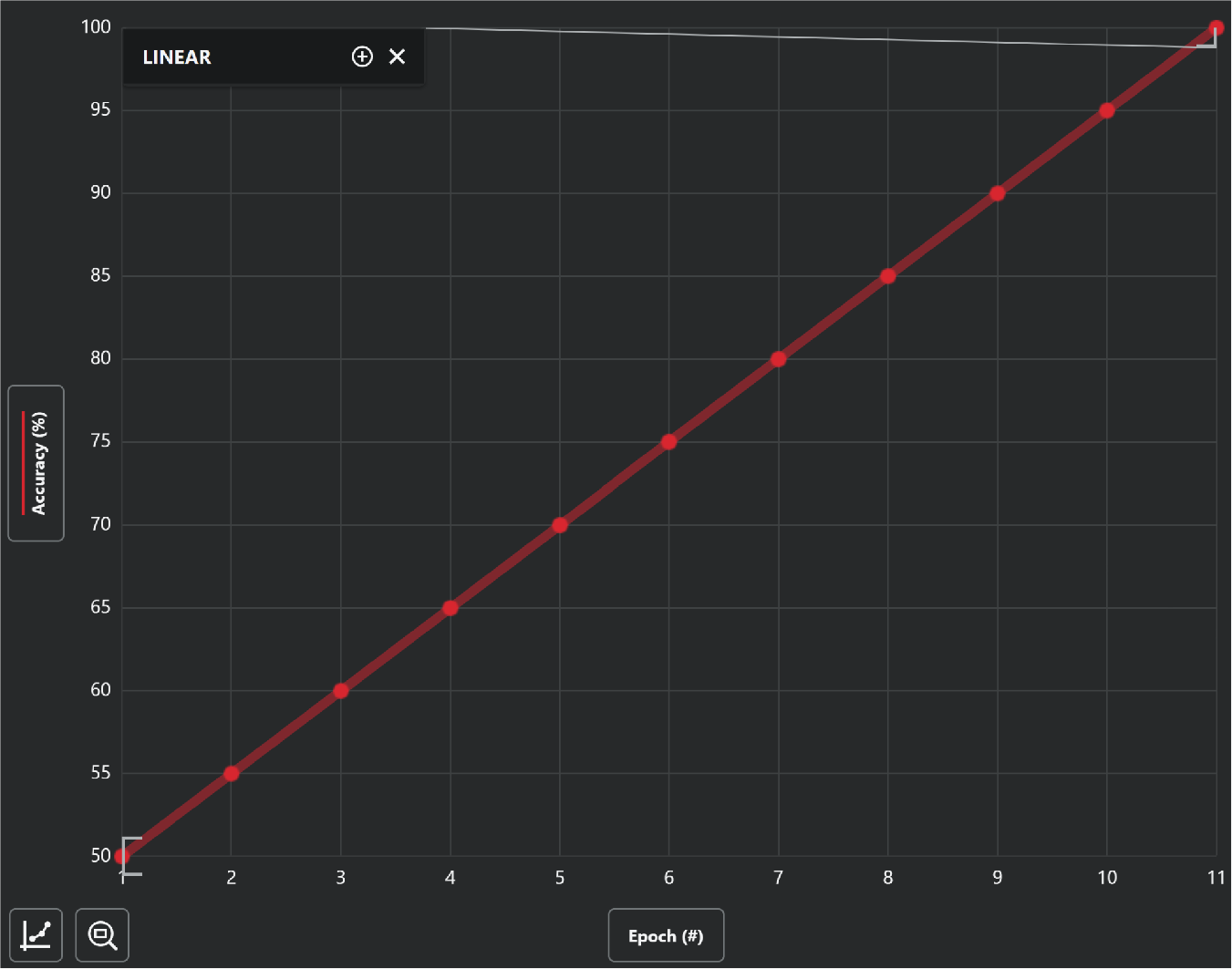
Linear graph showing increasing accuracy with every epoch passed. Epoch (#) × accuracyEpoch (#) × accuracy (%)

### 2.2 Construction of Tumor Classification Module

#### 2.2.1 Data Curation and Cleaning for the Tumor Classification Module

The data curation and cleaning process for the tumor classification module played a pivotal role in ensuring the reliability and efficacy of the deep learning system. We used 2 directories from the Cancer Imaging Archive; these 2 directories resulted in the model gaining access to 310 total images being split into roughly 100 images of each of the three major types of brain carcinoma (Metastasis, Glioma, and Meningiomas). The MRI images, sourced from the cancer imaging archive, underwent a meticulous quality assessment to address potential challenges. This included a thorough examination of image resolution to maintain consistency, noise reduction techniques to enhance clarity, and verification of a uniform imaging modality to prevent variations. We also grayscaled the images to filter out any image deformities present in the MRI images using OpenCV. We then cropped the extreme: top, left, bottom, and right portions of the scan; this was done to only include parts of the brain excluding blank spaces for a more accurate model. To better understand the cropping process, please refer to *Figure 2*. The inherent challenge of bodily movements during MRI sessions was addressed through an image registration process, aligning tumor regions across multiple images. An anomaly detection system was implemented to identify and rectify corrupted images or artifacts automatically. Labeling consistency was a focal point, with manual verification of tumor annotations to guarantee accuracy. Data augmentation techniques, such as rotation and flipping, were applied to diversify the training set and enhance the model’s generalization capabilities. Ethical considerations were paramount, with all data handling procedures aligning with privacy regulations and patient confidentiality standards. Looking ahead, continuous efforts will be made to expand the dataset, incorporating a broader range of patient demographics and tumor characteristics, ensuring the model’s adaptability to evolving medical scenarios.

#### 2.2.2 CNN Model Creation

The creation of the classification model is largely similar to the construction of the identification module. We used a convolutional neural network to create the tumor classification module. We again used a sequential model to base the module off of. The model begins with an input layer specifying an input shape of (220,220, 3). The model then adds two convolutional layers, each followed by a max pooling layer. The first convoluted layer has 85 filters with a kernel size of (3,3), with “valid padding” and ReLU activation for enhanced predictions. The second convoluted layer consists of 60 convoluted layers with similar settings. These 2 convoluted layers are designed to extract and analyze features of images for predictions. This is followed by dropout layers with a 25% drop rate; this is done to prevent a common problem such as overfitting. Then an output layer using sigmoid activation is compiled using the Adam optimizer and binary cross-entropy loss.

#### 2.2.3 Model Training

The classification model was trained in the same processes as the identification model. The training phase of the deep learning model uses the Keras library. It is training the previously defined sequential model on a dataset. The ‘model. fit’ method is used to train the model. It takes several arguments, including the training data (’x_train_scaled’ and ‘y_train’) and the number of training epochs, which is set to 17 in this case. During each epoch, the model will iteratively update its internal parameters to improve its performance on the training data. The ‘validation_data’ argument is set to a tuple containing the validation data (’x_test_scaled’ and ‘y_test’), allowing the model’s performance to be evaluated on a separate dataset during training to monitor its generalization ability.

#### 2.2.3 Model Testing

Following the training, the model is asked to evaluate its prediction accuracy for what images it believes to be either “Glioma”, “Metastatic”, or “Meningioma”. Instead of printing out each specific image, this time the model assigns a number to each MRI scan being fed; this is done to improve the execution time as the epoch number is doubled. When the model assessed itself, it achieved an accuracy of 91% and leaned toward correctly classifying images as “Glioma” and “Metastatic” as opposed to “Meningioma”; taking 100 seconds and 867ms. To combat the slight bias, we increased the learning rate from 10^™2^to 10^™4^. This would enhance the evaluated accuracy to 98%; taking 117 seconds and 652ms. A graph depicting accuracy with epoch number is shown in *Figure 6*.

**Figure 6:**
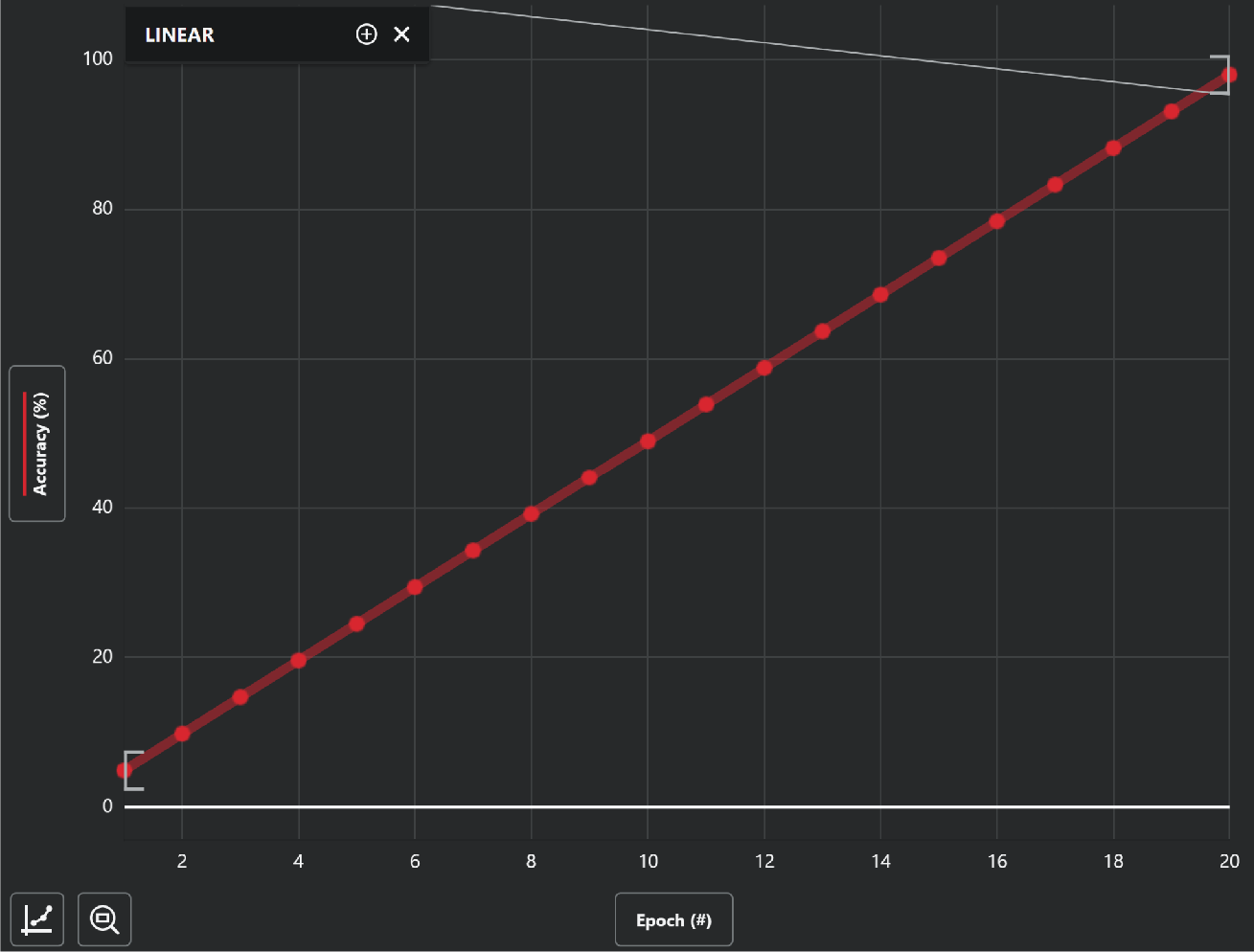
Linear graph showing increasing accuracy with every epoch passed. Epoch (#) × accuracy (%).

The slope was 4.9. Precision is found to be 97 using equation (1) below, recall is found to be 95 using equation (2) below, and the F-measure score is found to be 93.9, with equation (3).

1. *precision* = 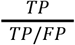
2. *recall or sensitivity* = 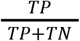
3. F − *measure score* = 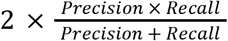

## 3.0 Discussion

In conclusion, this research presents a novel deep learning system leveraging Convolutional Neural Networks (CNNs) for the early detection and real-time classification of malignant brain tumors. The study addresses the challenges posed by the aggressive nature of classifying tumors during Medical Resonance Imaging (MRI). The proposed system demonstrates promising results in both tumor identification and tumor classification. The image classification module, trained on a carefully curated dataset from the Eindhoven University of Technology, achieved an accuracy of 97% after optimization. This accuracy is crucial for the early detection of tumors, offering a reliable tool for medical professionals to identify and intervene promptly. The constructed CNN model follows a sequential architecture designed for image classification tasks, incorporating convolutional layers for feature extraction and fully connected layers for decision-making. The training process involved careful curation of the dataset, model creation, and iterative training with the Keras library. The results indicate the model’s ability to learn and generalize effectively.

Overall, the proposed deep learning system shows promise in revolutionizing the detection and classification of malignant brain tumors. With the achieved high accuracy in image classification and substantial progress in tumor classification, this system holds the potential to significantly enhance the efficiency of targeted radiotherapy and improve patient outcomes. As the research suggests, further optimization and testing will contribute to refining the system and potentially achieving even higher levels of accuracy in tumor classification and medical imaging. Parts of this article’s readability were enhanced with the use of artificial intelligence LLMs. Future iterations will involve testing in real hospital systems.

## Data Availability

All data produced in the present study are available upon reasonable request to the authors
All data produced in the present work are contained in the manuscript

## Notes

### Competing Interest Statement

The authors have declared no competing interest.

### Funding Statement

This study did not receive any funding

### Author Declarations

The study used ONLY openly available human data that were originally located at the cancer imaging archive.

